# Interobserver Variability in Target Definition for Stereotactic Arrhythmia Radioablation

**DOI:** 10.1101/2023.03.01.23286657

**Authors:** Martijn H. van der Ree, Phillip S. Cuculich, Marcel van Herk, Geoffrey D. Hugo, Jippe C. Balt, Matthew Bates, Gordon Ho, Etienne Pruvot, Claudia Herrera-Siklody, Wiert F. Hoeksema, Justin Lee, Michael S. Lloyd, Michiel Kemme, Frederic Sacher, Romain Tixier, Brian V. Balgobind, Clifford G. Robinson, Coen R.N. Rasch, Pieter G. Postema

**Affiliations:** Amsterdam UMC location University of Amsterdam, Department of Cardiology, Meibergdreef 9, Amsterdam, the Netherlands; Amsterdam Cardiovascular Sciences, Heart Failure and arrhythmias, Amsterdam, the Netherlands; Department of Internal Medicine, Cardiovascular Division, Washington University School of Medicine, St. Louis, Missouri, USA; Department of Radiation Oncology, University of Manchester, Manchester Academic Health Centre, Manchester, UK; Department of Radiation Oncology, Washington University School of Medicine, St. Louis, Missouri, USA; Department of Cardiology, St. Antonius Hospital, Nieuwegein, the Netherlands; Department of Cardiology, South Tees Hospitals NHS Foundation Trust, Middleborough, UK; Department of Medicine, Division of Cardiology Cardiac Electrophysiology, Cardiovascular Institute, University of California San Diego, San Diego, California, USA; Heart and Vessel Department, Service of Cardiology, Lausanne University Hospital and University of Lausanne, Lausanne, Switzerland; Department of Immunity, Infection and Cardiovascular Disease, University of Sheffield Sheffield, UK; Section of Cardiac Electrophysiology, Emory University, Atlanta, Georgia, USA; Cardiac arrhythmia department, IHU LIRYC, Bordeaux University Hospital, Bordeaux, France; Department of Radiation Oncology, Amsterdam UMC, Amsterdam, the Netherlands; Department of Radiation Oncology, LUMC, Leiden, the Netherlands

**Keywords:** cardiac radioablation, stereotactic arrhythmia radiotherapy, stereotactic arrhythmia radioablation, ventricular tachycardia, interobserver variability

## Abstract

**Background:** Stereotactic arrhythmia radioablation (STAR) is emerging as a potential new therapy for patients with refractory ventricular tachycardia (VT). The arrhythmogenic substrate (target) is synthesized from clinical and electro-anatomical information. This study was designed to evaluate the baseline interobserver variability in target delineation for STAR.

**Methods:** Delineation software designed for research purposes was used. The study was split into three phases. Firstly, electrophysiologists (observers) delineated a well-defined structure in three patients (spinal canal). Secondly, observers delineated the arrhythmogenic cardiac VT target in three patients previously treated with STAR based on case descriptions. To evaluate baseline performance, a basic workflow approach was used, no advanced techniques were allowed (e.g. image integration). Thirdly, observers delineated three predefined segments from the cardiac 17-segment model. Interobserver variability was evaluated by assessing volumes, variation in distance to the median volume as expressed by the root-mean-square of the observer standard deviation (RMS SD) over the target volume, and the Dice coefficient.

**Results:** Ten electrophysiologists completed the study. For the first phase (spinal canal delineation), interobserver variability was low as indicated by low variation in distance to the median volume (RMS SD range: 0.02-0.02cm) and high Dice coefficients (mean: 0.97±0.01). In the second phase (VT-target delineation), distance to the median volume was large (RMS SD range: 0.52-1.02cm) and the Dice coefficients low (mean: 0.40±0.15). In the third phase (segment delineation), similar results were observed (RMS SD range: 0.51-1.55cm, Dice coefficient mean: 0.31±0.21)

**Conclusions:** Interobserver variability is high for manual delineation of the VT-target and ventricular segments. Difficulties in cardiac anatomical orientation on traditional radiation oncology CT scans appear to be an important driver of variability. This evaluation of the baseline observer variation shows that there is a need for methods and tools to improve variability and allows for future comparison of interventions aiming to reduce observer variation.

## BACKGROUND

Ventricular tachycardia (VT) is a life-threatening cardiac arrhythmia that is associated with increased risk of mortality and morbidity. Implantation of an implantable cardioverter defibrillator (ICD) reduces mortality, but ICD therapies are accompanied with their own adverse outcomes.^1,2^ In a subset of patients, the current state of the art, namely medication and radiofrequency ablation, fails. In these therapy-refractory patients, stereotactic arrhythmia radioablation (STAR, =cardiac radioablation) has been suggested as a bail-out procedure.^3^ In STAR, the ventricular arrhythmogenic substrate is treated by applying ionizing radiation. STAR is associated with a relatively durable reduction in VT episodes and a mostly mild acute toxicity profile in patients in patients during the first year.^3-10^ Longer term follow-up is currently accumulating.

With STAR, the arrhythmogenic substrate is first determined by aggregating clinical and electro-anatomical information to delineate a target substrate, upon which a 4D-CT scan is acquired for radiotherapy treatment planning purposes.^11^ Variables include a 12-lead electrocardiograms during VT if present, electroanatomical data from prior VT ablation(s), and cardiac imaging such as echocardiography, cardiac computed tomography imaging (CT), cardiac magnetic resonance imaging (CMR), and myocardial perfusion scintigraphy or F18-FDG positron emission tomography. In contrast to traditional planning for malignant tumors, the pro-arrhythmic substrate is not directly visualized on the radiotherapy planning 4D-CT scan, which complicates target definition and delineation. Thus, delineation is based on subjective and collaborative synthesis of the aggregate of many variables, which is naturally prone to large interobserver variability.^12,13^

The interobserver variability for target delineation in malignant tumors has been well quantified using established methods.^14^ Reducing observer variability allows for more standardized treatment, and, potentially, improved outcomes and reduced toxicity.^15^

To standardize and improve STAR treatment, the magnitude of interobserver variation in the context of STAR should be explored. This study was designed as a baseline study to evaluate and explore interobserver variation in target delineation for STAR. This baseline study will allow for future benchmarking of interventions aimed at reducing interobserver variation.

## METHODS

### Delineation and observers

Observers were required to have experience in the treatment of VT and of refractory VT using STAR. The observer panel was composed from electrophysiologists from different hospitals in Europe and the United States and observers were asked to perform contour delineation in three study phases.

To evaluate and explore the baseline interobserver variation in STAR, this study was split into three phases. Phase one consisted of delineation of a simple and well-defined anatomical structure: the spinal canal. This phase was used to verify whether the observers were able to delineate a well-defined structure using the study delineation software. In phase two, observers were asked to delineate the clinical target volume (CTV, VT-target without any additional uncertainty margins) for cardiac radioablation in three previously treated patients (see below: “Patients and CT-scans”) based on case descriptions with clinical and electro-anatomical information (text and images) as is common practice in STAR. Delineation instructions for the second phase included rules that are outlined in Table 1. To serve as a baseline study, a basic workflow approach was used and no advanced techniques such as resampling of the images in cardiology-preferred views and/or auto-segmentation of scans according to the AHA 17-segmented model or image integration were allowed.^13,16,17^ The third phase consisted of delineation of 3 predefined segments from the 17-segmented model (1 segment for each patient); basal-anterior (segment 1), mid inferoseptal (segment 9) and apical lateral (segment 16).^18^ This phase allowed exploration of interobserver variability in delineation of a predefined cardiac structure in conventional oriented scans (oriented to the body axes as traditionally used in radiation oncology, instead of the cardiac axes as traditionally used in cardiology). The delineation instructions and case descriptions are provided in the supplementary material.

**Table 1.** Rules for delineation of the clinical target volume in study phase 2.

| Rule |  |
| --- | --- |
| 1 | Choose scar or border zone, not healthy tissue |
| 2 | Choose only scar or border zone near VT exit sites, not necessarily the entire scar |
| 3 | Choose a single larger area, not multiple small areas |
| 4 | Goldilocks principle:<br>• Delineate too small, and you might miss the VT circuit<br>• Delineate too large, and there is (likely) higher risk for normal tissue injury |

### Patients and CT-scans

Phase 1 of the study included three patients who previously underwent an invasive catheter ablation for VT in the Amsterdam UMC and had a CT scan of the (Patient A, B and C). To enhance the visibility and delineation of the spinal canal, slices comprising of only one vertebra were selected allowing the spinal canal to be completely surrounded by bony structure. For phase 2 and 3 of the study, three patients suffering from therapy-refractory VT from the Washington University School of Medicine (St. Louis, Missouri, United States of America) previously treated with STAR were selected by the treating cardiologist (Patient 1, 2 and 3). Prior to STAR treatment, these patients underwent a 4D-CT scan for radiotherapy treatment planning purposes according to the local protocol (free breathing, patient in supine position with arms raised above the head in a forearm support, contrast enhancement when patient characteristics allowed and 1.5mm slice thickness). To reduce delineation times, slices with odd instance numbers were removed for phase 2 and 3. Outcome in terms of VT burden and safety were not part of this study. The institutional ethical review boards approved the study and patients gave written informed consent.

### Delineation tools

Specific delineation software, previously described in detail and specifically designed for research purposes, was used.^14^ This software includes delineation tools that are included in most commercial radiotherapy planning systems, but also enables detailed analyses of the delineation process. In the software, axial slices, and coronal and sagittal reconstruction of the CT scans were available for delineation. Per study phase, the observers received personal passwords to delineate and edit contours within the software. Only after completion of a phase, the observers received a new password for the subsequent phase. Observers were only able to view their own delineations.

### Contour analysis

Interobserver variability was evaluated by performing contour analysis. For every case in the three phases of the study, the delineated volumes were calculated per observer. The median surface of the delineated contours was computed in 3D, representing the 50% coverage of the contours (meaning that every voxel inside this median surface is delineated by ≥50% of the observers).^19^ The variation in perpendicular distance from this median volume to each observer’s individual contour was calculated. Distances larger than 20mm were set to 20mm. Then for each median volume point the standard deviation (SD) was calculated (and visualized) and the overall observer SD was expressed by the root-mean-square of the values in all points. The generalized Dice coefficient was calculated, which is a measure for overlap in volumes (Figure 1.).^20^ The Dice coefficient is expressed from 0-1 with 0 indicating no overlap in volumes, whilst a value of 1 indicates complete overlap. The Dice coefficient was calculated for all combinations of observers and then averaged. Data are presented as mean±standard deviation unless otherwise indicated.

**Figure 1.**
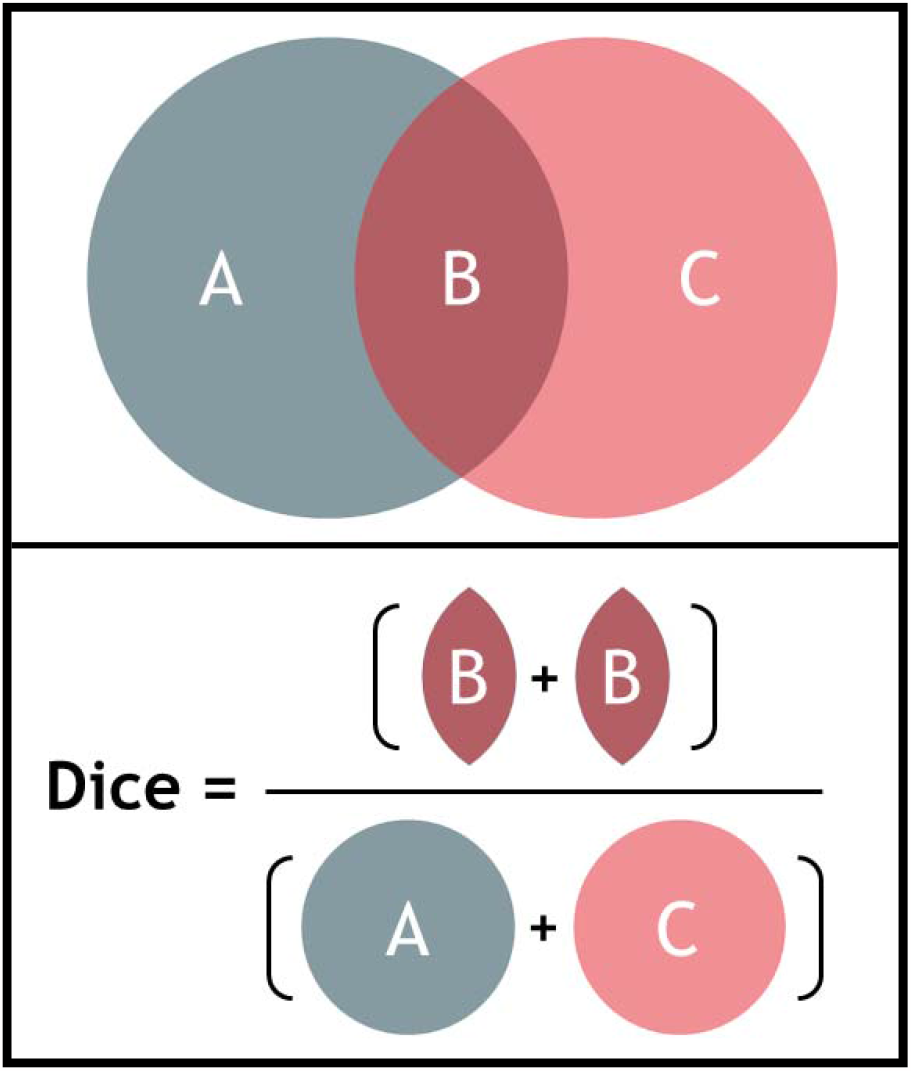
Illustration of the calculation of the Dice coefficient for two volumes A and C, B indicating the overlap. The Dice coefficient is calculated by dividing two times the volume of overlap (B) by the individual volumes (A+C) (formula: 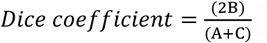).

## RESULTS

Ten observers from seven tertiary care hospitals completed the study. Table 2 shows the results for the three phases of the study. From figure 2, delineations from representative cases for each study phase can be appreciated. In figure 3 the standard deviation is projected on the median surface of the delineations for representative cases in each phase of the study.

**Table 2.** Results of the three study phases.

| Phase 1: Spinal canal |  |  |  |  |  |
| --- | --- | --- | --- | --- | --- |
|  | Median structure volume* (cm<sup>3</sup>) | Mean volume (cm<sup>3</sup>) | Range in volume (cm<sup>3</sup>) | RMS SD (cm) | Dice coefficient |
| Patient A | 1.78 | 1.56±0.03 | 1.52 - 1.62; 0.1 | 0.02 | 0.97±0.01 |
| Patient B | 2.97 | 2.58±0.06 | 2.47 - 2.67; 0.2 | 0.02 | 0.98±0.01 |
| Patient C | 2.08 | 1.79±0.04 | 1.7 - 1.82; 0.12 | 0.02 | 0.97±0.01 |
| Phase 2: VT-target |  |  |  |  |  |
|  | Median structure Volume* (cm<sup>3</sup>) | Mean Volume (cm<sup>3</sup>) | Range in volume (cm<sup>3</sup>) | RMS SD (cm) | Dice coefficient |
| Patient 1 | 25.8 | 34.6±17.4 | 9.9 - 61.4 | 1.02 | 0.32±0.17 |
| Patient 2 | 23.2 | 28.2±11.9 | 11.1 - 56.0 | 0.53 | 0.47±0.11 |
| Patient 3 | 16.9 | 22.7±7.8 | 10.8 - 36.7 | 0.52 | 0.41±0.11 |
| Phase 3: Segments |  |  |  |  |  |
|  | Median structure Volume *(cm<sup>3</sup>) | Mean Volume (cm<sup>3</sup>) | Range in volume (cm<sup>3</sup>) | RMS SD (cm) | Dice coefficient |
| Patient 1 | 6.3 | 13.5±9.5 | 6.4 - 35.2 | 1.55 | 0.21±0.19 |
| Patient 2 | 11.1 | 12.8±4.1 | 5.4 - 17.3 | 0.51 | 0.46±0.14 |
| Patient 3 | 5.4 | 9.4±4.7 | 3.2 - 19.9 | 0.55 | 0.25±0.20 |
| *The median structure volume represents the 50% coverage of the delineations, meaning that each voxel inside the median structure is delineated by at least 50% of the observers.<br>RMS SD: root-mean-square of the standard deviation |  |  |  |  |  |

**Figure 2:**
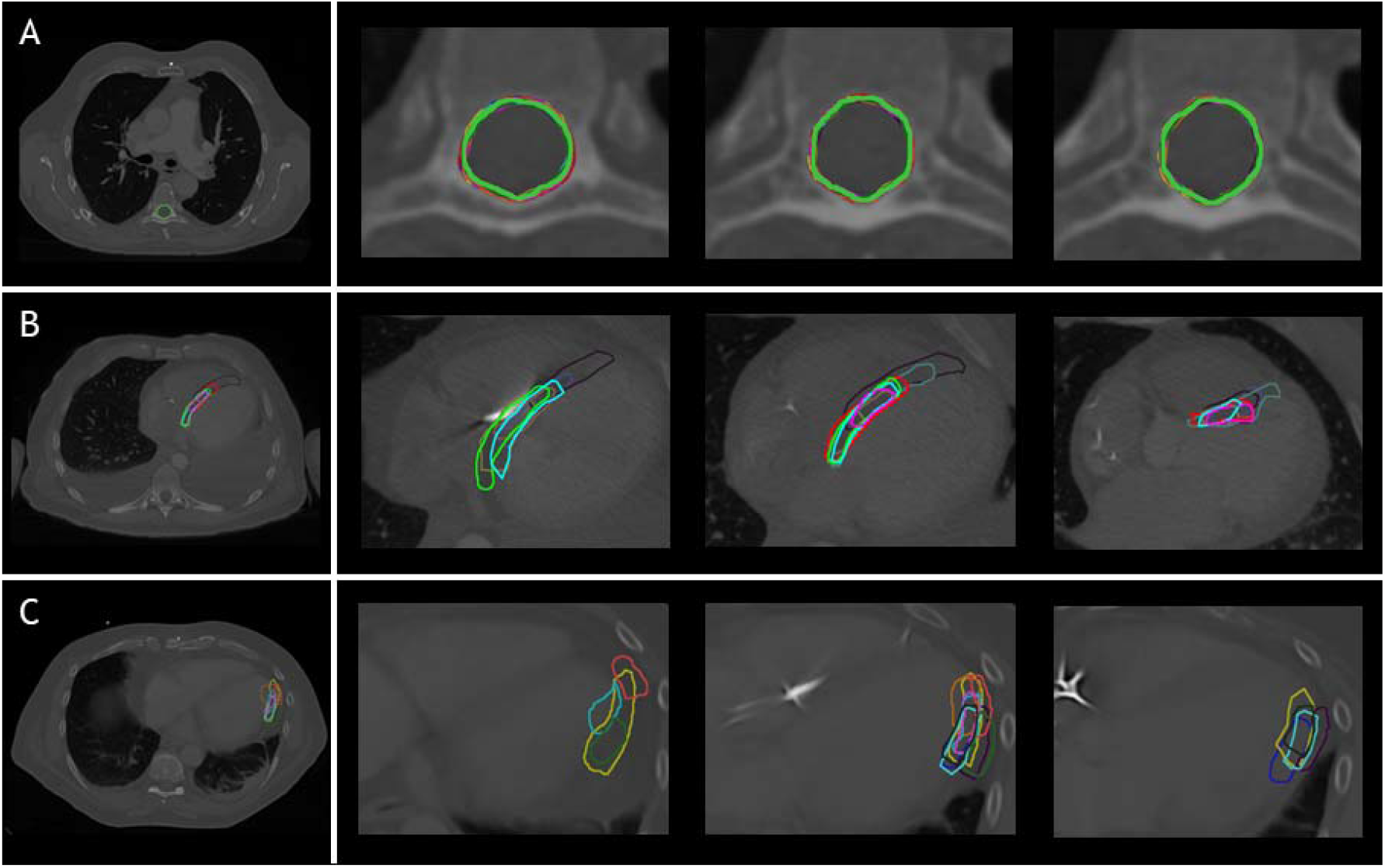
delineations in representative cases by the different observers (A) Phase 1: Spinal canal, patient A (B) Phase 2: VT-target, patient2 (C) Phase 3: Segments, patient 3 (segment 16: apical-lateral). Each color indicates a different observer.

**Figure 3:**
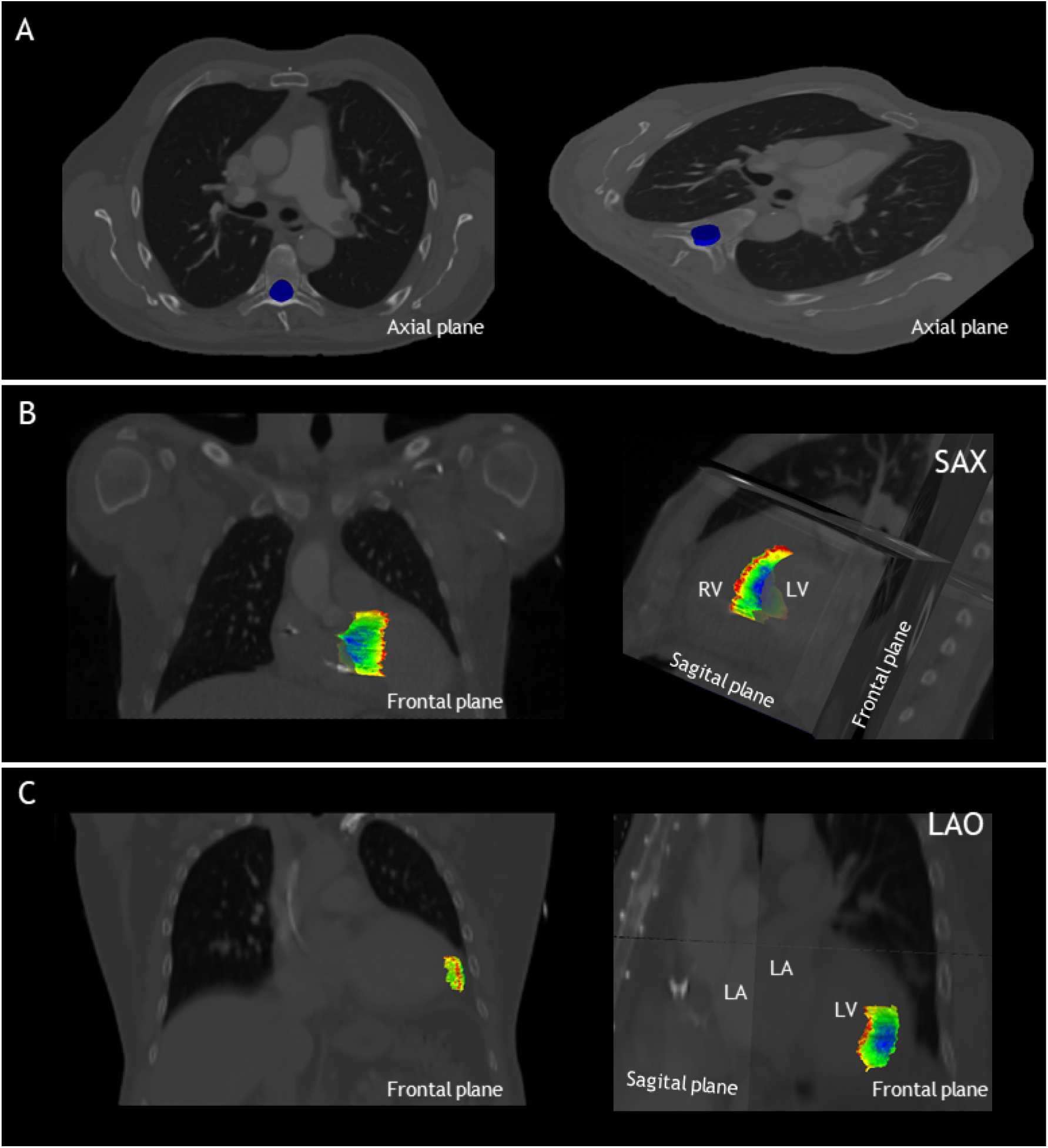
**T**he local standard deviation projected on the median structure of the delineations, from dark blue: SD<0.1 cm to red SD>1.5 cm. (A) Results for phase 1, delineation of the spinal canal, projected on the axial plane for patient A. Note that the standard deviation is below <0.1cm for the entire spinal canal. (B) Results for phase 2 of patient 2, delineation of the VT-target, projected on the frontal and sagittal planes. (C) Results for phase 3 of patient 3, delineation of the apical-lateral segment, projected on the frontal and sagittal plane. LA: left atrium, LAO: left anterior oblique view, LV: left ventricle, RV: right ventricle and SAX: cardiac short-axis view.

### Phase 1: Spinal canal

Low interobserver variation was found for the first phase of the study, the delineation of the spinal canal. This is indicated by low variation in the volumes (table 2), low variation in distance to the median volume (RMS SD range 0.02-0.02cm), and very high Dice coefficients (mean: 0.97±0.01) (table 2, figure 2A and 3A).

### Phase 2: VT-target

#### Patient characteristics

Patient characteristics of the 3 test cases for phase 2 included patients of age 54-70 years, all non-ischemic cardiomyopathy and left ventricular ejection fraction between 20-32%. Treatment included anti-arrhythmic drugs consisting of both amiodarone and mexiletine and previous catheter ablations (n≥4 per patient). For targeting, 12-lead VT ECGs, noninvasive electrocardiographic imaging (VT-exit site), echocardiography, CMR and nuclear imaging were available in all patients. In two (67%) patients, no recent electroanatomical maps from invasive catheter ablations were available and in one (33%) patient the diagnostic cardiac CT-scan was not available.

#### Results

Delineation of the VT-target based on case descriptions without advanced techniques resulted in high interobserver variability (table 2, figure 2B and 3B). The mean target volume ranged from 23-35 cm^3^ and widely differed per observer (range: 9.9–61.4 cm^3^, table 2), as did the variation in distance to the median volume (RMS SD range: 0.5-1.02cm, table 2). The mean Dice coefficient for all three patients was 0.40±0.15 with a range of 0.32-0.47 indicating low volume overlap.

### Phase 3: Segments

For phase 3, delineations of segments from the 17-segmented model showed equally high interobserver variation (table 2, figure 2C & 3C) as indicated by differences in delineated volumes between observers (table 2), large variation in distance to the median volume (RMS SD range: 0.51-1.55cm) and Dice coefficients below 0.5 (mean: 0.31±0.21)

## DISCUSSION

In this baseline study, the interobserver variation in target volume delineation for STAR was explored. Firstly, we observed that cardiologist-electrophysiologists can delineate a well-defined anatomical structure with low interobserver variability in software specifically designed for this purpose. Secondly, for delineation of the VT-target using a basic workflow approach, interobserver variability was high. Lastly, interobserver variability was also high in delineation of predefined segments from the 17-segmented heart model.^18^

Our findings are congruent with previous studies evaluating interobserver variation in delineation of the VT-target for STAR treatment.^12,13^ In these studies the VT-target delineations were compared to consensus delineations^12^ or based on a head-head comparison of two observers.^13^ This study adds to the prior data in its demonstration of interobserver variation in the context of STAR treatment among an experienced and intercontinental (Europe and America) group of observers basing our results on median volumes and comparisons for each combination of observers. Noteworthy, when comparing the different studies, it is important to acknowledge that this study was designed as a baseline study allowing to evaluate interventions to reduce observer variation in the future.

An important aspect of this analysis is our use of a control arm in phase 1, which shows the generally good agreement among our observer cohort for identifying basic radiologic borders. It supports the conclusion that variation in subsequent phases cannot be explained by difficulties in the delineation process itself, but are due to other sources of uncertainty, e.g. including challenging cardiac anatomic orientation in conventional radiation oncology CT scans oriented to the long-axis of the body (instead of following cardiac axes).

The high interobserver variation for VT-target delineation we observed in phase 2, does not necessarily mean there is no consensus on the pro-arrhythmic regions In general, delineations did show overlap (figure 2B). As can be appreciated from figure 3B, there appears to be consensus on the core of the pro-arrhythmic substrate. Importantly, during conventional ablation electrophysiologists interact with the substrate, and the effect of applying ablative energy can be directly observed. This allows for a better understanding of the pathophysiology and the localization of the VT-substrate underlying the ventricular arrhythmias and probably reduces variability due to direct feedback. It is important to acknowledge that in this study, observers did not have an interaction with the substrate and their delineations were merely based on text and images presented in the case descriptions. Although this is a limitation, such a workflow is also part of STAR treatment, because delineation is always separately performed and does not include direct feedback. Moreover, when patients referred for STAR treatment recently underwent a (high-risk) procedure in another hospital or are not able to undergo a (repeat) invasive catheter ablation due to limiting patient characteristics (e.g. insufficient access), an off-line aggregation of the electro-anatomical data (preferably together with the principal operator of the last VT-ablation) will be performed. The fact that Dice coefficients in phase 3 (predefined segments) was similar as in phase 2 (VT-target) could indicate that the interpretation of electroanatomical information is not the main driver of the interobserver variation. Instead, difficulties in cardiac anatomical orientation on traditional radiation oncology CT-scans appears to be an important driver of variability (i.e. no contrast enhancement and orientations to the long-axis of the body). This is supported by the fact that in patient 1, in which the CT-scan was not contrast enhanced due to the patient’s renal insufficiency, the variability was particularly high in both phase 2 and 3.

Indeed, in the third phase of the study, as mentioned above, we also found high interobserver variability.^18^ This is likely explained by the fact that these segments are defined based on a cardiac orientation and not on the conventional orientation with planes perpendicular to the long-axis of the body as used in radiotherapy planning systems. While the 17-segmented model as structured approach interpretation and delineation of the VT-target has previously been proposed,^16^ we here show that manual delineation in scans not angulated to the cardiac orientation leads to undesirable results.

### Potential interventions to reduce interobserver variation

Now that the baseline interobserver variation in the context of STAR is determined, future research should focus on methods to reduce this variability. Several methods and techniques are already used in clinics worldwide. ^13,16,17,21^ Currently, the best strategy to reduce interobserver variation and improve efficacy and safety needs to be decipher.

#### Reorientation and segmentation

Firstly, cardiologist-electrophysiologists are used to the cardiac orientation in imaging. We therefore believe that re-orienting and re-sampling the images could ease delineation and reduce interobserver variation. As previously mentioned, the 17-segmented model could be used to enhance orientation in cardiac anatomy.^16^ Based on our results and previous work, re-orientation and segmentation should be performed in a (semi-)automated manner as this has been shown to be reproducible.^17^

#### Delineation teams

Currently, STAR treatment is only used in patients with refractory VT. As a result, eligible patients are highly complex. Peer review of targets has demonstrated to increase contour agreement in radiation oncology.^22^ Potentially, by discussing and delineating cases with cardiologist-electrophysiologists (or in multi-disciplinary teams) a reduction in interobserver variability could be achieved as well.

#### Image integration

VT-target delineation is based on results of several electroanatomical modalities. Integrating all the different modalities into radiotherapy planning systems could result in lower interobserver variability. However, matching imaging data with electro-anatomical maps from previously performed catheter ablation will introduce new uncertainties and matching errors. This notwithstanding, efforts are being undertaken to develop robust methods for image integration.^21,23,24^

### Clinical implications

High interobserver variation in STAR treatment is undesirable as this leads to differences in treatments between centers. It is unknown whether the variation observed results in differences in treatment plans or clinical outcomes, although this would be conceivable. The interobserver variation may seem clinically very high. However, when interpreting these results, it is important to consider that also for the current state-of-the-art treatments, e.g. VT ablation, differences between operators and hospitals exist (for example, due to experience of the operator, diagnostic work-up and different ablation techniques and strategies).^1,2^ These differences, however, have not been quantified. In contrast, in radiation oncology there is a long history of great interest in interobserver variability in target delineation that has led to international harmonization and standardization. Therefore, we believe our results are not discouraging, but should rather be seen as a starting point to improve and standardize STAR treatment to improve efficacy and safety.

Given the large observer variation one may wonder why the clinical outcomes of STAR are so good and it is possible that this is due the nature of the disease: contrary to a tumor it may be that STAR is effective if part of the substrate is irradiated.

## CONCLUSION

The interobserver variation in the context of STAR treatment was high for delineation of the VT-target using a basic workflow approach. Difficulties in cardiac anatomical orientation on traditional radiation oncology CT scans appear to be an important driver of variability. To standardize STAR treatment, future studies should focus on interventions aiming to reduce this variability. The results of this baseline evaluation will allow for future comparisons of such interventions.

## Data Availability

The study data are available from the corresponding author on reasonable request.

## ACKNOWLEDGEMENTS

None.

## SOURCES OF FUNDING

Dutch Heart Foundation grant 03-003-2021-T061 to Dr. Postema. This project has been supported by the Foundation “De Drie Lichten” in The Netherlands.

## DISCLOSURES

Washington University receives research support from Varian Medical Systems, Siemens Healthineers, Mevion, and ViewRay. GDH: Consulting for Varian Medical Systems. GH: Equity in Vektor Medical Inc and consulting for Abbott and Kestra. JL: Consulting for Varian Medical Systems. CGR: Consulting for Varian Medical Systems.

## Notes

### Clinical Trial

Not applicable.

### Author Declarations

The institutional ethical review boards (Amsterdam UMC, Washington University) approved the study and patients gave written informed consent.

